# Estimation of the basic reproduction number of COVID-19 from the incubation period distribution

**DOI:** 10.1101/2021.11.04.21265937

**Authors:** Lasko Basnarkov, Igor Tomovski, Florin Avram

## Abstract

**Background:** The estimates of future course of spreading of the SARS-CoV-2 virus are frequently based on Markovian models in which the transitions between the compartments are exponentially distributed. Specifically, the basic reproduction number *R*_0_ is also determined from formulae where it is related to the parameters of such models. The observations show that the start of infectivity of an individual appears nearly at the same time as the onset of symptoms, while the distribution of the incubation period is not an exponential.

**Methods:** We propose a method for estimation of *R*_0_ for COVID-19 based on the empirical incubation period distribution and assumed very short infectivity period that lasts only few days around the onset of symptoms. It is tested on daily new cases in six major countries in Europe, in the first wave of epidemic in spring, 2020.

**Results:** The calculations show that even if the infectivity starts two days before the onset of symptoms and stops immediately when they appear, the value of *R*_0_ is larger than that from the classical, Markovian approach. For more realistic cases, when only individuals with mild symptoms spread the virus for few days after onset of symptoms, the respective values are even larger.

**Conclusions:** The calculations of *R*_0_ and other characteristics of spreading of COVID-19 based on the classical, Markovian approaches should be taken very cautiously. Instead, non-Markovian models with general distribution functions of transition between compartments should be considered as more appropriate.

**Key messages:**

- Although formulae for estimate of the basic reproduction number *R*_0_, by using general-form functions of infectivity are known since the earliest works in epidemiology, majority of studies are based on exponential distribution function.
- We introduce a new methodology of calculating *R*_0_ with an infectivity function obtained by combining empirical incubation period distribution with infectivity window function that is localized around the onset of symptoms.
- Estimates of *R*_0_ for the first wave of COVID - 19 in the spring 2020, by the proposed methodology are larger than those from the classical SIR model.
- When possible, the estimates of *R*_0_ should be based on empirical distributions of the infectivity functions, while the values obtained with the conventional epidemic spreading models should be taken with caution.

## 1 Introduction

The mathematical epidemiology is a field of study with increasing public importance by providing valuable insights for the authorities aimed for planning various actions against epidemic diseases. Although its inception dates back to Daniel Bernoulli [1], the major development as separate field of study might be attributed to the works of Ross [2–4] and Kermack and McKendrick [5]. The key quantity that is usually reported from studies about epidemic spreading is the basic reproduction number (or rate, or ratio) *R*_0_. Its importance is particularly emphasized with the fact that the herd immunity, achieved naturally or through vaccination expressed as fractions of the total population depends on it as 1 *−* 1*/R*_0_ [6]. The basic reproduction ratio represents an estimated number of newly infected persons by one infected individual introduced in a completely susceptible population. In its calculations, even in the earliest works it was considered that the infectivity potential of the infector depends on the time passed since she or he become infected - the age of infection, adequately modeled with certain function with general form. However, in majority of works the dominant role has exponential function *e*^*−rt*^. This choice provides simple relationships for the basic reproduction ratio, but also makes the epidemic spreading models to have simple form based on ordinary differential equations. This exponential function in the probabilistic approach has a meaning of probability density function of the period in which individuals stay in given compartment (exposed, infected, and so on), before transiting to another one. This is the appropriate function when the Markovian property holds, which means that the probability for transition to another state is independent on the past. However, very often this assumption is not empirically verified, and particularly for the diseases like the COVID-19, which can have particularly long period of incubation [7–9]. This practically means that the spreading potential of an infected individual becomes significant only when the incubation period is near its end. As a consequence, more appropriate, and more accurate estimates would be obtained by non-Markovian approach which allows for using arbitrary distribution functions of the incubation period duration.

Since the COVID-19 has become serious threat of the health and seriously disrupted the normal life, large number of studies have been made for modeling its spread and particularly about estimates of the basic reproduction ratio. It is calculated on a daily basis by various institutions and individuals. We have an impression that majority of the methods for its estimate are based on relationships that result from compartmental models with ordinary differential equations, that is, with the Markovian setting and the related exponential distribution of incubation period. It is thus our motivation to provide an estimate of *R*_0_ with an approach that is based on the non-exponential distribution of the incubation period, in a non-Markovian framework, which we believe is particularly relevant for the spread of the SARS-CoV-2. A basis of our calculations are the empirically obtained distributions of the time to onset of symptoms. Such distributions are further combined with certain window function that assumes that an individual is infectious only few days around the onset of symptoms. The combination is an infectivity function that determines the basic reproduction number. The obtained results for different European countries in the first wave of the epidemic, when the virus was assumed to spread freely, are larger than those with the classical, Markovian approach. The key outcome of these observations is that the estimates of *R*_0_ with the classical models should be considered very carefully. Finally, it is worth noting that the method used here has also similarities with the early studies in demography by Richard Böckh [10] and in epidemiology by Alfred Lotka [11].

The paper is organized as follows. In the Section 2 we first explain the formula used for estimation of *R*_0_. In the following Section 3 it is elaborated on the infectivity function that is used for estimates of *R*_0_. In the next, Section 4 are presented the results and a discussion on them and we finish the paper with the Conclusions.

## 2 Methods

The derivation of the formula that we use for estimation of the basic reproduction rate *R*_0_ for general infectivity function can be found in various works in the literature (for example in [12–14]). For completeness we present it here and chose to use a discrete-time approach since it is more appropriate for the available data. To start with, assume that the epidemic is in inception phase which means that the number of newly infected individuals grows exponentially. As is the case of COVID-19, when daily new cases are reported, by denoting with *I*_*d*_(*t*) the confirmed cases for day *t* given as fraction of the total population, one has the following exponential form

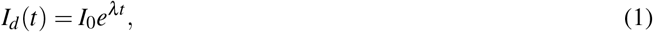

where *λ* is a parameter for the growth rate, while *I*_0_ is a constant. The growth rate *λ* is related with the period of doubling with *T*_*d*_ = ln 2*/λ*. For many diseases the individuals are not able to infect the others immediately, but after certain period has passed. We consider that the infectivity potential of the individuals that have contracted the spreading agent at the same moment *t*, is described with certain infectivity function *i*(*τ*). We assume that this function depends on the time elapsed since the individuals have become exposed, but not on the moment when that happened and has finite support *T* which means that *i*(*τ*) = 0, for *τ > T*. The shape of this function will be elaborated later on. Denote with *I*(*t, τ*) the fraction of infectees that have become infected at moment *t*, by having a contact with infectors that have become infected *τ* time units earlier. The function *I*(*t, τ*) depends on the fraction of susceptibles, but also on the fraction of those previously infected *I*_*d*_(*t − τ*) and on their infectivity potential encoded in *i*(*τ*). Thus one has the following form

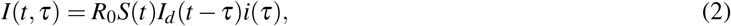

where the constant of proportionality *R*_0_ will be shown to be exactly the basic reproduction ratio. The fraction of the new infectees will be obtained as sum of contributions from all possible infectors

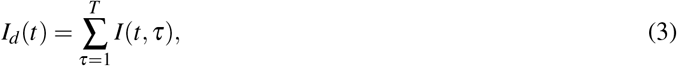

which will further result in the following recurrent relationship

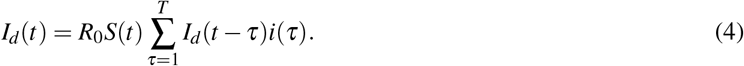

By applying the exponential form for of the function of newly infected individuals (1), in the last relationship, one will have

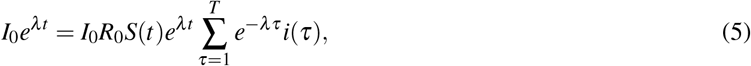

from where it follows that

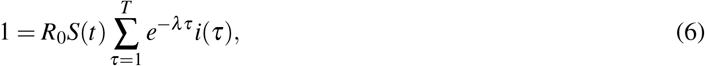

Since at the inception of the epidemic *S*(*t*) *≈* 1, one has the reproduction number

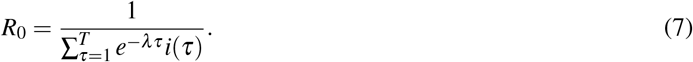

If one has obtained *R*_0_ from the last expression and it is assumed to be the same during the epidemic, than the herd immunity threshold *S*_th_, which corresponds to unity reproduction number *R*, can be obtained from (6) as

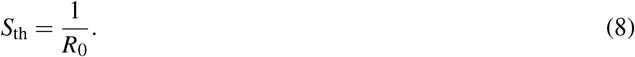

### 2.1 Self-consistency relationship for *R*_0_

The basic reproduction number *R*_0_ represents an estimated multiplicative factor determining the number of newly infected individuals that will contract the spreading agent in a contact with certain infected individual. Here we use it to denote the growth factor that corresponds to the group of individuals that have been exposed in the same period. Thus, form one side, the fraction of all infectees that have contracted the pathogen from infectors that become infected at certain moment *t* should be *R*_0_*I*_*d*_(*t*). From another side, this fraction can be represented as forward-time sum as follows

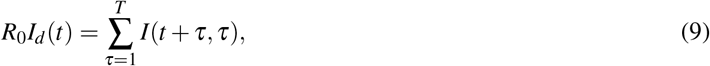

which accounts for all admissible combinations *I*(*t* + *τ, τ*). By using (2), one has

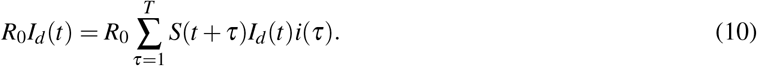

For slowly growing epidemic one might assume that the fraction of susceptibles does not change significantly in the considered period *S*(*t*) *≈* 1. Then, one can see that the constant *R*_0_ in the relationship (2) will be the basic reproduction number, if the infectivity function is normalized 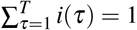.

### 2.2 Continuous-time formula

Although analysis of the data in this work is based on discrete time, for completeness we provide the continuous-time version, that can be found in the literature in similar forms. Denote with *I*(*t*) the fraction of the population that has become infected within infinitesimal interval (*t, t* + *dt*), and assume that it grows exponentially at the onset of epidemics *I*(*t*) = *I*_0_*e*^*λt*^. These infectees have appeared from contacts with others that have been infected in the past *I*(*t − τ*). Again, let the function *R*_0_*i*(*τ*) denote the infectivity potential at some later moment *τ* of the group of individuals infected nearly at the same time. As for the discrete-time case assume that *i*(*τ*) has some finite support (0, *T*). Now, the formula for newly infected population will read

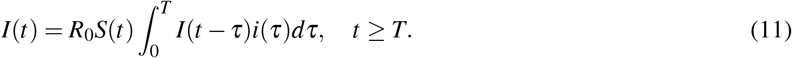

One should note that similar relationship has appeared earlier in the works of Ross and Hudson [15]. By plugging in the exponential form of the newly infected individuals one will obtain

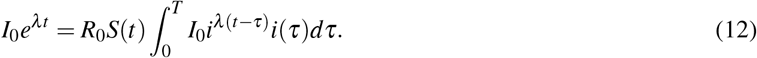

which will reduce to similar relationship as the discrete-time one

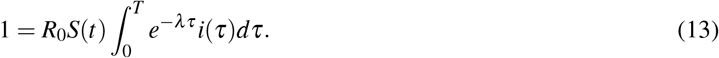

At the onset of an epidemic *S*(*t*) *≈* 1, the reproduction number will be

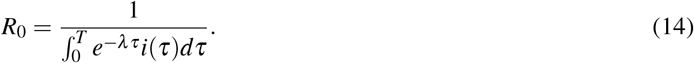

In reality, one does not have observations of the short-interval infections *I*(*t*), but for certain unit interval, like a day. So, one has to choose a function *I*(*t*) such that its daily integral 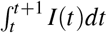 would fit the observed data.

## 3 Shape of the infectivity function

The function *i*(*τ*) should model the infectivity potential of the fraction of the population, becoming infected within the same unit interval, at some later moment *τ* after contracting the spreading agent. Its shape can be directly deduced from epidemiological tracing of infector-infectee pairs. Our approach is to relate it with other characteristics of the respective disease. First, its shape depends on the fractions of the individuals that have not been recovered yet and thus is related to the function that describes the healing process. However, since the COVID-19 is a disease which has long period of recovery we consider that the function modeling it can be considered as constant for the period under study. Second, the infectivity function depends on the incubation period distribution, for which one can find many studies in the literature. Third, the viral load in reality is not constant over time, which means that during infectiousness, a person cannot spread the pathogen with equal intensity [16]. For simplicity in this work it is also considered to be constant. As a side note we mention here that the product of the healing and incubation period functions was applied in non-Markovian epidemic spreading models recently [17, 18], while the same relationship with other functions was used in the pioneering work of demography [10]. Thus, in this work we are focusing on using the distribution of the time to onset of symptoms as basis. To reach the final shape of the infectivity function we combine it with an infectivity window function *w*(*t*). It is non-zero in the period *t ∈* [*t*_init_; *t*_end_], between certain initial moment *t*_init_ before the onset of symptoms, and ends at *t*_end_ after. From practical perspective, the shape of the window in the segment [*t ∈ t*_init_, 0], with *t* = 0 being the time of onset of symptoms, may be obtained from sensitivity analysis conducted on known infector-infectee pairs, as suggested in [16]. The shape of *w*(*t*) for *t ∈* [0; *t*_end_] represents the distribution of time from onset of symptoms to self isolation, quarantining or viral clearance, obtained from epidemiological data. In our approach we assume that after the onset of symptoms, a person with more severe symptoms would immediately reduce the contacts with the others and thus would not be a significant infector any more. Those with mild symptoms, that constitute about 80% of the cases [19], would continue with their normal daily life and would spread the virus until *t*_end_, when they receive positive test results. Thus the infectivity window function has a shape of two steps

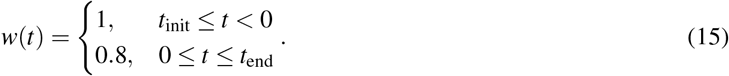

The infectivity function was considered to be the convolution of the incubation period distribution *β* (*τ*) and the infectivity window *w*(*τ*)

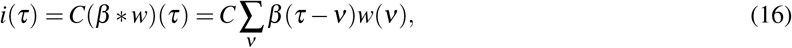

where *C* is a constant determined from the normalization condition 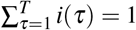.

We compare our calculations with those obtained from the classical (Markovian) SIR model where the basic reproduction ratio with the growth rate *λ* is related with [13]

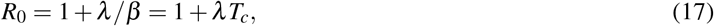

where *β* is the infectivity rate, and *T*_*c*_ is the mean infectivity period – the average value of the duration of the exponentially distributed infective state.

## 4 Results and discussion

We have used the data from the Our World in Data database for the numbers of daily cases in the first wave in the spring 2020 for six major European countries. As a data window for study we have taken the period from the first day when every day new cases were reported until the moment when lockdown measures were introduced. This choice was made under the assumption that in that period the virus SARS-CoV-2 was spreading nearly freely in the population, with only positive cases being isolated. From those numbers we have fit an exponential curve and estimated the exponential growth rate factor *λ*. The results are summarized in the table 1.

**Table 1:** Countries under study and the estimated exponential growth factor

| Country | Start date | End date | $\lambda$ |
| --- | --- | --- | --- |
| Germany | February 25 | March 12 | 0.298 |
| France | February 25 | March 11 | 0.322 |
| Italy | February 21 | March 9 | 0.235 |
| Russia | March 12 | March 30 | 0.171 |
| Spain | February 25 | March 11 | 0.343 |
| United Kingdom | February 23 | March 22 | 0.237 |

We have used the estimates of the growth factor *λ* in the expression for the calculation of the basic reproduction number (7). The infectivity function *i*(*τ*) was obtained from the convolution (16) of windows with variable width, and incubation period functions from two sources in the literature [7, 8].

The onset of infectiousness was considered to be two [16] or one day before the appearance of symptoms. We have also assumed that after the onset of symptoms only those with mild symptoms, that are about 80% [19] can be considered as further infectors. As the end of the infectiousness period was considered either the same day or two days after the onset of the symptoms. The first case is very conservative and is related to the assumption that all individuals would be very cautious in their contacts with the others and are not infectors at all. The second one is less conservative and likely a more realistic scenario, where those with mild symptoms do not change their daily routines and spread the virus freely before receiving positive test. Finally, the results from the classical SIR model are also provided for comparison. Details about the considered scenarios is given in table 2.

**Table 2:**
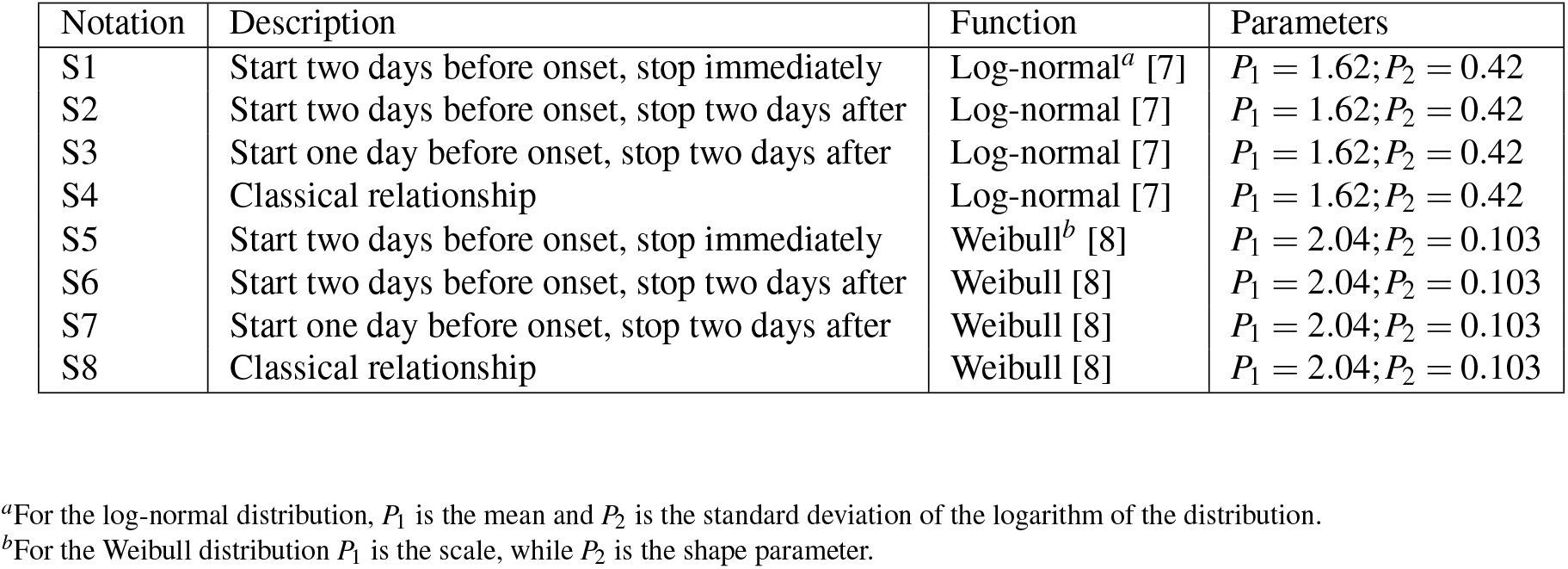
Description of different scenarios for calculation of basic reproduction ratio

| Notation | Description | Function | Parameters |
| --- | --- | --- | --- |
| S1 | Start two days before onset, stop immediately | Log-normal<sup>a</sup> [7] | $P_1 = 1.62; P_2 = 0.42$ |
| S2 | Start two days before onset, stop two days after | Log-normal [7] | $P_1 = 1.62; P_2 = 0.42$ |
| S3 | Start one day before onset, stop two days after | Log-normal [7] | $P_1 = 1.62; P_2 = 0.42$ |
| S4 | Classical relationship | Log-normal [7] | $P_1 = 1.62; P_2 = 0.42$ |
| S5 | Start two days before onset, stop immediately | Weibull<sup>b</sup> [8] | $P_1 = 2.04; P_2 = 0.103$ |
| S6 | Start two days before onset, stop two days after | Weibull [8] | $P_1 = 2.04; P_2 = 0.103$ |
| S7 | Start one day before onset, stop two days after | Weibull [8] | $P_1 = 2.04; P_2 = 0.103$ |
| S8 | Classical relationship | Weibull [8] | $P_1 = 2.04; P_2 = 0.103$ |

<sup>a</sup>For the log-normal distribution, $P_1$ is the mean and $P_2$ is the standard deviation of the logarithm of the distribution.
<sup>b</sup>For the Weibull distribution $P_1$ is the scale, while $P_2$ is the shape parameter.

The estimates of *R*_0_ for the different scenarios for all six European countries are presented in table 3. One can easily note that in all scenarios the values of *R*_0_ from the non-Markovian approach considered here are larger than those from the classical, Markovian one. In the overly restricted scenarios S1 and S5 when onset of symptoms, even for those with mild symptoms, is a reason for immediate self-quarantine leads to larger estimates than the reference cases S4, and S8, respectively. The more realistic assumptions in the other cases produce even larger estimates. In the reality there would have been cases for even longer period of infectivity than two days after the onset of symptoms, and this could be the true for significant fraction of infected population. Although we have not tried to model this, the logic suggests that the longer the infectivity period, after the onset of symptoms, the larger *R*_0_ is. This implies that the values of *R*_0_ based on the classical Markovian SIR model for the COVID-19 are likely significant underestimates. From this observation one has that the more accurate estimate of *R*_0_ needs knowledge of the infectivity window function *w*(*τ*), that is the period when an infected person is infectious, in relation to the onset of symptoms. The situation can be even more complicated if the infectivity window function is dependent on the incubation period duration. It could be possible for those that develop symptoms earlier to correspond one infectivity window function, while for those that develop them after longer period another one. Thus, besides being very cautious with using the estimated *R*_0_ from the classical approach, it is needed to have good estimate of the infectivity function in order to have more correct value of the basic reproduction ratio.

**Table 3:** Estimated basic reproduction ratio *R*_0_ for six European countries for the eight different scenarios.

| Country | S1 | S2 | S3 | S4 | S5 | S6 | S7 | S8 |
| --- | --- | --- | --- | --- | --- | --- | --- | --- |
| France | 3.22 | 4.06 | 5.02 | 2.87 | 5.37 | 6.72 | 8.01 | 3.50 |
| Germany | 3.00 | 3.73 | 4.53 | 2.73 | 4.93 | 6.10 | 7.17 | 3.31 |
| Italy | 2.45 | 2.95 | 3.42 | 2.37 | 3.85 | 4.60 | 5.22 | 2.83 |
| Russia | 1.97 | 2.27 | 2.51 | 1.99 | 2.87 | 3.29 | 3.60 | 2.33 |
| Spain | 3.43 | 4.38 | 5.49 | 2.99 | 5.78 | 7.31 | 8.81 | 3.66 |
| UK | 2.47 | 2.97 | 3.45 | 2.38 | 3.89 | 4.65 | 5.28 | 2.84 |

## 5 Conclusions

We have estimated the basic reproduction ratio *R*_0_ by using the more general non-Markovian framework, that besides being known from the emergence of mathematical epidemiology, has not been widely applied. The approach was used to determine the value of *R*_0_ in six major countries in Europe during the first wave of the COVID-19 epidemic. The onset of infectiousness, instead of starting immediately after contraction of the pathogen, was taken to be related to the onset of symptoms, for which the empirical evidence suggests that is not distributed exponentially as the Markovian assumption implies. The incubation period distribution was further combined with an infectiousness window function which was considered to have short period - one or two days before and finish up to two days after the onset of symptoms. From both functions we have constructed an infectivity function that uniquely determines *R*_0_. In all scenarios we have considered, the calculated value for *R*_0_ was obtained to exceed the one from the classical relationship. This suggests that the calculations with the classical, Markovian approach, should be taken rather cautiously.

Better estimates of *R*_0_ would be obtained with empirical function of the infectiousness window, or direct estimation of the shape of the infectivity function. This needs more involving epidemiological tracing, that is not an easy task. However, we hope that the observation that the classically obtained value of *R*_0_ is an underestimate would lead to more intensive work for gathering epidemiological data and increase the awareness that non-Markovian setting in the epidemic models should be given more attention.

## Data Availability

All data produced in the present study are available upon reasonable request to the authors

## 7 Author contributions statement

L.B. and I.T. initiated the study of non-Markovian approach for estimation of the basic reproduction number, formulated the basic model and made the numerical calculations. L.B., I.T. and F.A. contributed to the final model and the overall form of the manuscript.

## 8 Acknowledgement

This research was partially supported by the Faculty of Computer Science and Engineering, at the Ss. Cyril and Methodius University in Skopje, Macedonia.

